# HPV prevalence, vaccination coverage and intention to get vaccinated among gay, bisexual, and other men who have sex with men: Evaluation of Quebec’s (Canada) HPV vaccination program

**DOI:** 10.64898/2026.05.13.26352734

**Authors:** Chantal Sauvageau, Alain Fourmigue, Manale Ouakki, Gilles Lambert, Ann N. Burchell, François Coutlée, Alexandra de Pokomandy, Marc Brisson, Troy Grennan, Milada Dvorakova, Daniel Grace, Darrell H. S. Tan, Trevor A. Hart, Joseph Cox

**Author notes:** **Corresponding author** Chantal Sauvageau, 2400 D’Estimauville Québec G1E 7G9. **Contributions** Chantal Sauvageau: Conceptualization, Funding acquisition, Formal analysis, Investigation, Methodology, Project administration, Resources, Supervision, Validation, Writing – original draft, Writing – review & editing Alain Fourmigue: Data curation, Formal analysis, Methodology, Software, Validation, Writing – original draft, Writing – review & editing Manale Ouakki: Conceptualization, Methodology, Validation, Writing – review & editing Gilles Lambert: Conceptualization, Funding acquisition, Methodology, Resources, Supervision, Validation, Writing – review & editing Ann Burchell: Conceptualization, Funding acquisition, Methodology, Resources, Supervision, Validation, Writing – review & editing François Coutlée: Conceptualization, Formal analysis, Investigation, Methodology, Resources, Validation, Writing – review & editing Alexandra de Pokomandy: Conceptualization, Methodology, Validation, Writing – review & editing Marc Brisson: Writing – review & editing Troy Grennan: Writing – review & editing Milada Dvorakova: Methodology, Validation, Writing – review & editing Daniel Grace: Methodology, Validation, Writing – review & editing Darrell H. S. Tan: Writing – review & editing Trevor A. Hart: Conceptualization, Funding acquisition, Methodology, Validation, Writing – review & editing Joseph Cox: Conceptualization, Funding acquisition, Methodology, Resources, Supervision, Validation, Writing – review & editing.

## Abstract

**Objectives:** In Quebec, Canada, vaccination against human papillomavirus (HPV) has been publicly-funded since January 2016 for gay, bisexual, and other men who have sex with men (GBM) aged ≤26 years. The study aimed to analyze data collected in Greater Montreal (Engage study) to evaluate the HPV vaccination program for GBM in Quebec.

**Study Design:** Engage is a cohort of sexually active GBM aged ≥16 recruited via respondent-driven-sampling (RDS) in Canada. Participants completed a questionnaire and tested for sexually transmitted infections.

**Methods:** RDS-II weights were applied to adjust for recruitment. Subgroups were compared using standardized mean differences. Odds ratios of HPV vaccination and prevalence ratios of anal HPV infection adjusted for potential confounders were estimated using robust regression models.

**Results:** Of 1179 participants, 309 were eligible for free HPV vaccination. Vaccine coverage among eligible GBM was 42%. Among those who disclosed same-sex sexual activity and discussed HPV vaccination with their healthcare provider, coverage reached 82%. Anal HPV prevalence among eligible GBM was 26.5% for ≥1 HPV-6/11/16/18 genotypes without significant difference between vaccinated and unvaccinated individuals. Among unvaccinated GBM aged ≤26 who were aware of the vaccine, 60% intended to get vaccinated within the next year.

**Conclusions:** One to two years after GBM aged ≤26 were included in the Quebec HPV vaccination program, 42% of eligible GBM in Greater Montreal had been vaccinated. Anal HPV prevalence was high among GBM. Vaccinees were more likely to self-report a prior STI diagnosis. Offering vaccination to all preadolescents in schools appears essential to maximize vaccination benefits.

## 1. INTRODUCTION

In Quebec, Canada, free human papillomavirus (HPV) vaccination has been available since January 2016 for gay, bisexual, and other men who have sex with men (GBM) aged ≤26[1], and since September 2016 for Grade 4 boys (9-10-years-old). Free HPV vaccination became available to immunocompromised individuals or people living with Human immunodeficiency virus (HIV) aged ≤26 in 2014 (≤45 since 2022)[1]. The quadrivalent vaccine (targeting HPV-6/11/16/18 genotypes) has been available for men through the private market since 2010[2]. The nonavalent vaccine replaced the quadrivalent in September 2016. Several reasons justified the inclusion of GBM in the program, including the high burden of HPV-related infections and lesions among GBM[3,4].

The prevalence of ano-genital HPV infections has been shown to be at least threefold higher among GBM than among heterosexual men[3–5]. HPV infections cause anogenital and oropharyngeal cancers[6,7]. Rates of anal cancers among GBM have been estimated to be 30-100 higher than those of the general male population[8]. In Quebec, approximately 66 new anal cancer cases are diagnosed annually, with approximately 90% caused by HPV, mostly HPV-16[3,6], making most anal cancers vaccine-preventable.

HPV vaccine clinical trials have shown high efficacy in males, preventing 84% of incident anal vaccine-type infections in GBM aged ≤26 who were HPV-naive and had ≤5 lifetime sexual partners[9]. Additionally, effectiveness against anal HPV in GBM is highest with early HPV vaccination before sexual activity debut[10]. Limited data are available on HPV vaccination acceptability and effectiveness in real-world settings among sexually active GBM who may have been HPV-exposed before vaccination[11,12].

The Engage study (https://www.engage-men.ca/) is a Canadian cohort initiated in 2017 in the greater regions of Montreal, Toronto and Vancouver among sexually active GBM aged ≥16. The study examined GBM’s sexual health, presenting an opportunity to evaluate Quebec’s vaccination program among GBM. Some HPV-related findings from the Engage study have been previously published[13–16]. In 2017-2019, HPV vaccination coverage (≥1 dose) was 26–35% for GBM aged ≤26[13]. Pooled anal HPV prevalence among GBM aged ≤30 was 66.7% (95%CI: 59.7–73.6) for all genotypes and 25.4% (95%CI: 19.5–31.3) for HPV-6/11/16/18 genotypes[14]. These papers did not report on the acceptability of discussing sexual practices with a healthcare provider to obtain free HPV vaccination and the age at which GBM would be comfortable doing so.

Using data from the Engage Study (Greater Montreal area), we aimed to evaluate the HPV vaccination program for GBM. Specifically, we: (1) assessed HPV vaccination coverage, characteristics of vaccinated and unvaccinated GBM, as well as factors associated with vaccination; (2) examined disclosure of same-sex sexual activity to healthcare professionals to obtain the HPV vaccine; (3) measured and compared anal HPV prevalence between vaccinated and unvaccinated GBM; and (4) evaluated intention to get vaccinated among unvaccinated GBM still eligible for free HPV vaccination.

## 2. MATERIALS AND METHODS

### 2.1. Participants, Recruitment and Data Collection

We used data from the Engage study that enrolled GBM in Greater Montreal, including the Island of Montreal, Laval, and Longueuil from February 2017 to June 2018. Participants were recruited via respondent-driven sampling (RDS), a chain-referral recruitment method designed for sampling hard-to-reach populations[17].

At enrollment, to be eligible, participants had to: 1) be aged ≥16, 2) identify as a cis- or transgender man and 3) have had recent sexual intercourse with another man (past 6 months).

Participants completed a questionnaire (Appendix A for the questions pertaining to key variables) and underwent sexually transmitted infections (STI) screening. Only participants aged ≤30 were tested for HPV infections to focus on GBM mainly targeted by free HPV vaccination program. Men self-collected anal specimens using a moistened Dacron swab. Research staff explained self-collection procedures and provided illustrated and validated instructions to participants[18].

### 2.2. Variables of interest

#### 2.1.1. Eligibility for Free HPV Vaccination

In Quebec, HPV vaccination has been offered free of charge to GBM aged ≤26 years since January 2016. For this analysis, we distinguished between participants who were eligible for free vaccination at program launch (age ≤26 years in 2016) and those who remained eligible at the time of enrollment (age ≤26 years at enrollment).

#### 2.2.2. HPV Vaccination Status

Participants self-reported their vaccination status and age at vaccination. Considering the study period, we assume that most participants received the quadrivalent vaccine[1]. HPV vaccination (receiving ≥1 dose) was treated as a binary variable. Participants were classified as unvaccinated if they answered “no” or “unsure” regarding their vaccination status, or if they were unaware of the HPV vaccine.

#### 2.2.3. Disclosure of same-sex sexual activity

Disclosure of same-sex sexual activity to healthcare professionals was examined among participants eligible for free HPV vaccination and who had a regular healthcare professional. When applicable, participants were asked their age at first disclosure. Responses <9 years were considered invalid and excluded.

#### 2.2.4. HPV Genotyping

HPV DNA was extracted from self-collected swab samples using the MasterPure DNA Purification Kit (Epicentre Biotechnologies) as described previously[15]. HPV identification was performed in two stages. First, all samples underwent a generic test to detect HPV DNA presence[19]. Second, HPV DNA-positive samples were genotyped using the Roche Linear Array HPV that identifies 36 HPV genotypes[20]. Samples reactive for HPV52 in the Linear Array were then analyzed using a validated HPV52-specific real-time PCR assay[21]. To verify sample validity, a β-globin DNA sequence was amplified, and samples were considered valid if β-globin and/or HPV DNA was detected.

High-risk oncogenic HPV genotypes were defined according to the International Agency for Research on Cancer classification[22].

#### 2.2.5. HPV Vaccination Intention

Intention to receive the HPV vaccine free of charge was assessed among unvaccinated participants aged ≤26, as they were still eligible for free vaccination at the time of enrollment. Intentions were dichotomized as “likely/very likely” versus “undecided/unlikely/very unlikely”.

### 2.3. Statistical Analysis

RDS recruitment is known to oversample individuals who are more visible in the community due to its chain-referral approach[17]. Weighting[23] is commonly used to adjust RDS data. For each participant, the propensity for inclusion in the sample, known as visibility score[24], was imputed and inverted to derive RDS-II weights, which were applied to produce population-level inferences about GBM.

Unadjusted proportions were calculated to characterize the full sample. Standardized mean differences (SMD) were used to compare participants, e.g. vaccinated vs unvaccinated, with thresholds for negligible (<0.2), small (0.2–0.5), moderate (0.6–0.8), and large (>0.8) differences[25].

Odds ratios (ORs) of vaccination (≥1 dose) were estimated by robust logistic regression with RDS-II weights. Collinearity among factors was assessed using variance inflation factor. Model selection used the Akaike information criterion to avoid overfitting.

Prevalence ratios (PRs) of HPV (6/11/16/18 types) between vaccinated (≥1 dose) and unvaccinated GBM were estimated using robust Poisson models with RDS-II weights. Potential confounders of the association between vaccination and HPV infection were identified through literature review and the authors’ expertise. Effective confounders showing imbalance (SMD>0.2) were included in the model regardless of statistical significance. Lifetime self-reported diagnosis of STI was additionally evaluated as an effect modifier by including an interaction term in the model, with stratified estimates presented to aid interpretation. Regression analyses were restricted to participants with complete data. Appendix B shows the participant flowchart outlining the number of participants included in each of the analyses.

All analyses were performed using R v4.1.2 (R Core Team, 2021).

### 2.4. Ethical approval

The Engage Montreal Study was approved by the research ethics board of the Research Institute of the McGill University Health Centre (#MP-CUSM-15-632).

## 3. RESULTS

### 3.1. Participants’ characteristics

Overall, 1179 participants were recruited in greater Montreal. Half (52.8%) identified as French Canadian, 20.9% were ≤26 years-old, 14.0% immigrated to Canada (last 4 years), 44.4% had university-level education, 41.9% earned <20 000 CAD annually, 81.8% identified as gay, 27.0% smoked daily, and 67.5% had a regular healthcare provider. Self-reported lifetime prevalence of STIs and condylomas was 55.0% and 23.8%, respectively. Laboratory-confirmed HIV infection was detected among 18.3% of participants.

### 3.2. HPV vaccine coverage

Of 1179 participants, 309 were eligible for free HPV vaccination at program launch (January 2016). Among these, 125 (40.4%) reported ≥1 dose of vaccine before enrollment. The RDS-adjusted estimate of vaccine coverage among GBM eligible for free HPV vaccination was 41.5% (95%CI 35.3-47.9). The median age at first dose was 23 years (IQR: 21-27 years), and 98% were vaccinated after first sexual intercourse (Figure 1).

**Figure 1.**
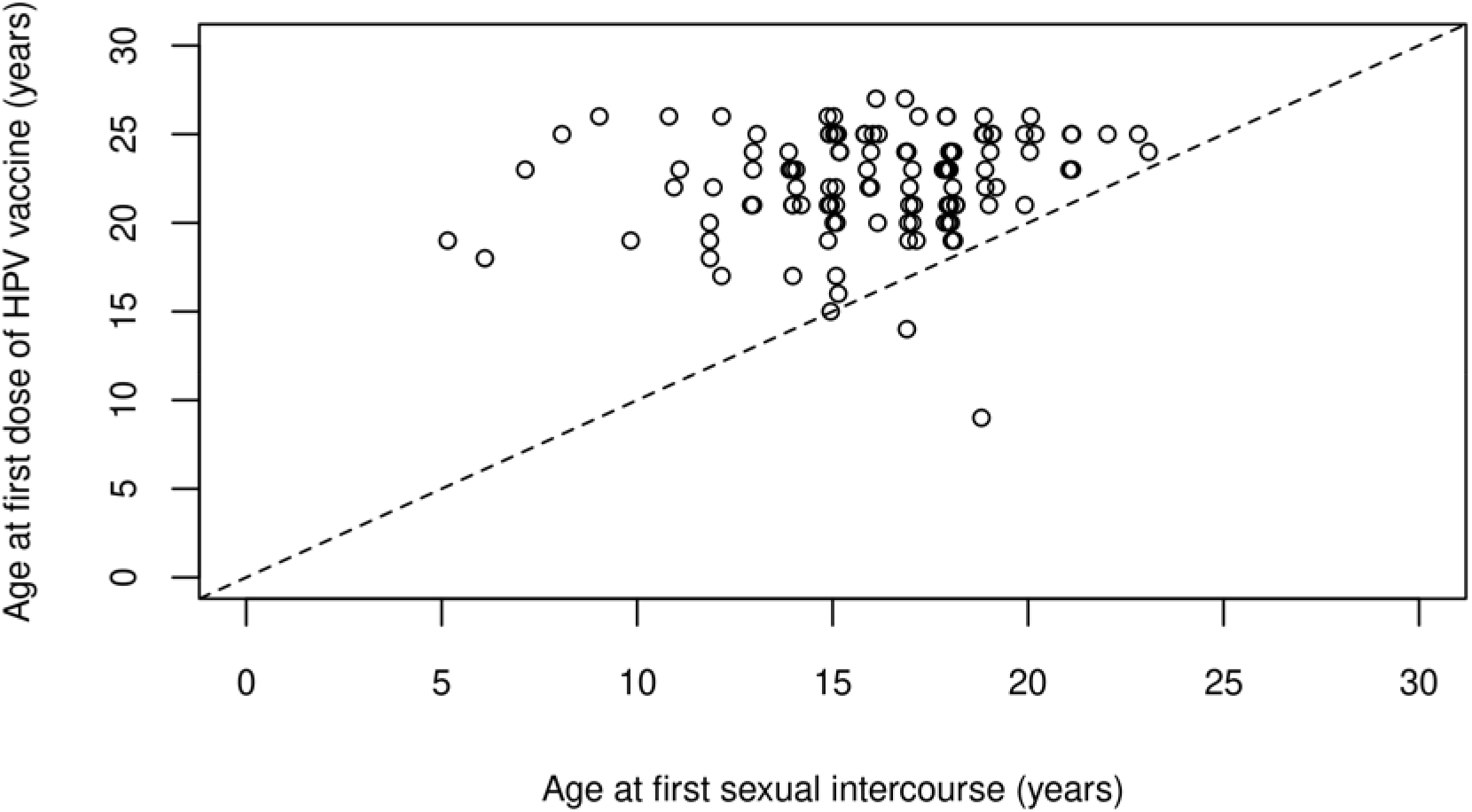
Age at first dose of HPV vaccine and age at first sexual intercourse among participants eligible for free HPV vaccination who reported having received ≥1 dose (n=125)

### 3.3. Differences between vaccinated and unvaccinated participants

The proportion of participants who reported having discussed HPV vaccination with a healthcare provider was 96.5% among vaccinated versus 18.3% among unvaccinated (SMD=2.4). The proportion of participants who knew that the HPV vaccine was recommended for GBM was 95.5% among vaccinated versus 45.5% among unvaccinated (SMD=1.4). Other differences with SMD>0.2 are shown in Table 1.

**Table 1.**
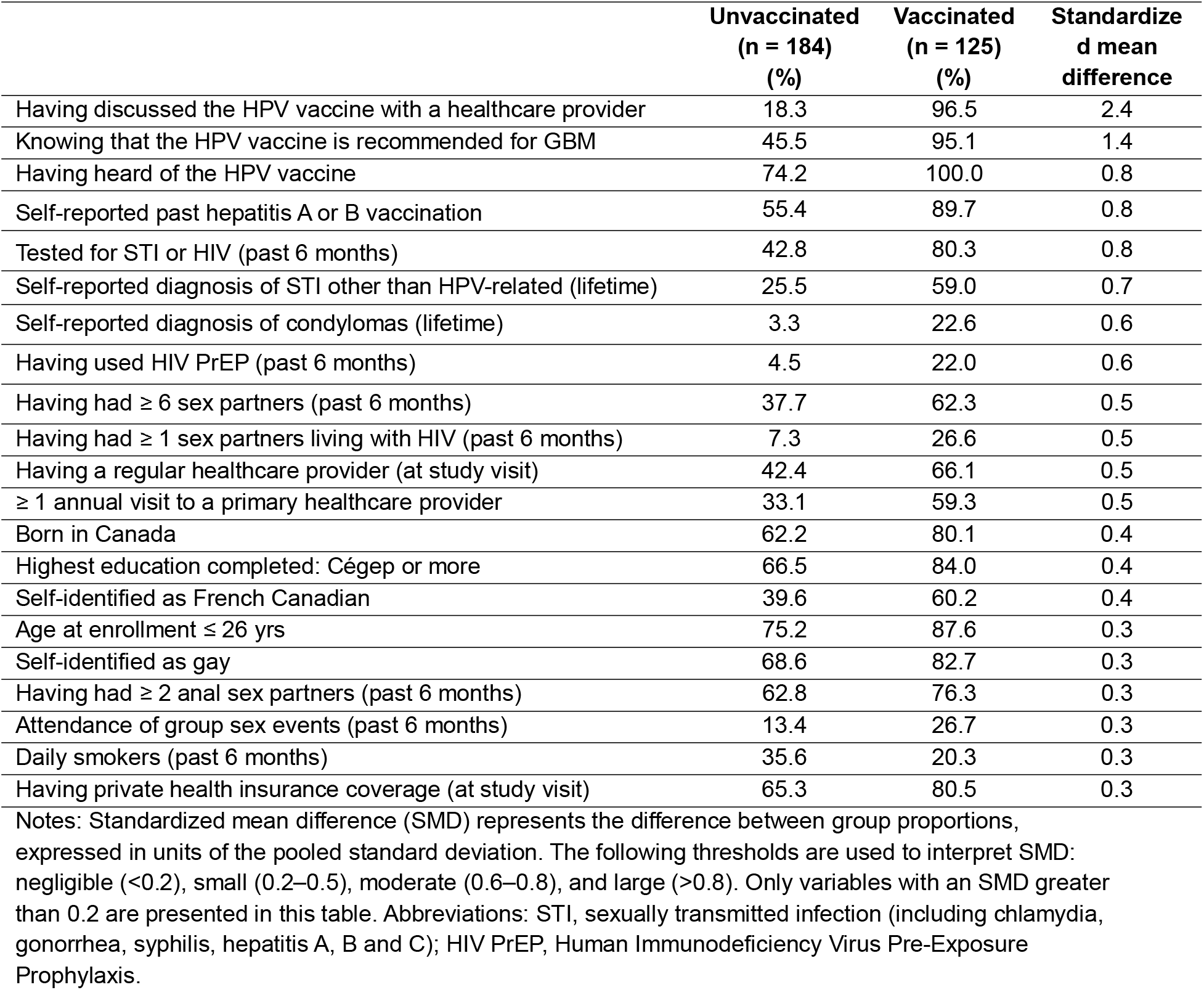
Largest differences between unvaccinated and vaccinated (≥1 dose) GBM eligible for free HPV vaccination (n=309)

### 3.4. Factors associated with HPV vaccination

Variables most strongly associated with HPV vaccination were a self-reported past hepatitis A or B vaccination (OR=7.5; 95%CI: 2.8–19.8) and a previous self-reported diagnosis of condylomas (OR=6.1; 95%CI: 1.9–19.4) (Table 2).

**Table 2.** Odds-ratios (OR) of vaccination (≥1 dose) among GBM eligible for free HPV vaccination (n=296)

| | Vaccinated ($\geq 1$ dose)<br>/ total participants | Univariable<br>regression | Multivariable<br>regression |
| --- | --- | --- | --- |

|  | Count | % | OR | 95%CI | OR | 95%CI |
| --- | --- | --- | --- | --- | --- | --- |
| Self-reported past hepatitis A or B vaccination |  |  |  |  |  |  |
| Never or don't remember | 15 / 88 | 17.0 | Ref |  | Ref |  |
| ≥ 1 dose | 108 / 208 | 51.9 | 6.9 | 3.5 - 13.7 | 7.5 | 2.8 - 19.8 |
| Self-reported diagnosis of condylomas (lifetime) |  |  |  |  |  |  |
| Never | 98 / 262 | 37.4 | Ref |  | Ref |  |
| ≥ once | 25 / 34 | 73.5 | 8.3 | 3.5 - 20.0 | 6.1 | 1.9 - 19.4 |
| Using HIV-PrEP (past 6 months) |  |  |  |  |  |  |
| No | 97 / 259 | 37.5 | Ref |  | Ref |  |
| Yes | 22 / 29 | 75.9 | 9.3 | 3.6 - 23.8 | 4.3 | 1.4 - 13.8 |
| Living with HIV | 4 / 8 | 50.0 | 2.4 | 0.6 - 10.5 | 0.9 | 0.1 - 6.4 |
| Tested for STI or HIV (past 6 months) |  |  |  |  |  |  |
| No | 27 / 126 | 21.4 | Ref |  | Ref |  |
| Yes | 96 / 170 | 56.5 | 5.9 | 3.3 - 10.6 | 3.3 | 1.5 - 7.1 |
| Age at enrollment |  |  |  |  |  |  |
| ≥ 27 yrs | 17 / 59 | 28.8 | Ref |  | Ref |  |
| ≤ 26 yrs | 106 / 237 | 44.7 | 2.3 | 1.1 - 4.7 | 3.3 | 1.5 - 7.4 |
| Education (highest level completed) |  |  |  |  |  |  |
| High school or less | 21 / 70 | 30.0 | Ref |  | Ref |  |
| Cégep (College) or more | 102 / 226 | 45.1 | 2.6 | 1.3 - 5.0 | 2.8 | 1.2 - 6.5 |
| Frequency of visits to primary health care provider |  |  |  |  |  |  |
| No care provider or < once a year | 51 / 165 | 30.9 | Ref |  | Ref |  |
| ≥ once per year | 72 / 131 | 55 | 2.9 | 1.7 - 5.1 | 2.4 | 1.2 - 4.7 |
| Self-reported diagnosis of STI other than HPV related (lifetime) |  |  |  |  |  |  |
| Never | 54 / 181 | 29.8 | Ref |  | Ref |  |
| ≥ once | 69 / 115 | 60.0 | 4.7 | 2.7 - 8.1 | 2.1 | 1.0 - 4.2 |
| Born in Canada |  |  |  |  |  |  |
| No | 24 / 92 | 26.1 | Ref |  | Ref |  |
| Yes | 99 / 204 | 48.5 | 2.3 | 1.2 - 4.2 | 2.1 | 1.0 - 4.5 |
| Number of sex partners (past 6 months) |  |  |  |  |  |  |
| ≤ 5 | 48 / 151 | 31.8 | Ref |  | Ref |  |
| > 5 | 75 / 145 | 51.7 | 2.9 | 1.7 - 4.9 | 1.6 | 0.8 - 3.2 |

Notes: Regression analysis performed on 296 (96%) participants with complete data. Abbreviations: STI, sexually transmitted infection (including chlamydia, gonorrhea, syphilis, hepatitis A, B and C); HIV PrEP, Human Immunodeficiency Virus Pre-Exposure Prophylaxis

### 3.5. Disclosure of same-sex sexual activity to obtain the HPV vaccine

Of 309 participants eligible for free HPV vaccination, 163 (52.7%) had a regular healthcare provider. Among these, 134 (82.2%) felt comfortable discussing sexual health issues with their healthcare provider, 124 (76.1%) had disclosed same-sex sexual activity, and 87 (53.4%) had discussed HPV vaccination. The median age at which participants first disclosed same-sex sexual activity to a healthcare provider was 19 years (IQR: 17-25 years) (Appendix C). Vaccine coverage among GBM eligible for free HPV vaccination who both disclosed same-sex sexual activity and discussed HPV vaccination with their healthcare provider was 81.9% (95%CI: 72.0-88.8)

### 3.6. Anal HPV Prevalence

Of 309 participants eligible for free HPV vaccination, 287 enrolled after HPV specimen collection began (delays occurred in starting this procedure) and provided a sample; 191 had valid HPV results. Anal HPV prevalence among GBM eligible for free vaccination was 71.2% (95%CI: 63.4-77.9) for ≥1 HPV genotypes and 26.5% (95%CI: 19.8–34.5) for ≥1 quadrivalent genotypes (HPV-6/11/16/18) (Table 3).

**Table 3.** Anal HPV prevalence among GBM eligible for free HPV vaccination (n=191)

| HPV Infections | Number of positive samples | Prevalence (%) | 95%CI |
| --- | --- | --- | --- |
| $\geq 1$ genotypes | 136 | 71.2 | 63.4 - 77.9 |
| Simple infection (only one HPV genotype) | 54 | 26.7 | 20.4 - 34.2 |
| Multiple infections (several HPV genotypes) | 82 | 44.5 | 36.7 - 52.5 |
| $\geq 1$ high-risk oncogenic genotypes | 87 | 46.1 | 38.3 - 54.1 |
| Other than high-risk oncogenic genotypes | 49 | 25.1 | 19.0 - 32.5 |
| $\geq 1$ quadrivalent genotypes (HPV-6/11/16/18) | 46 | 26.5 | 19.8 - 34.5 |
| Other than quadrivalent genotypes | 90 | 44.7 | 37.0 - 52.6 |
| $\geq 1$ nonavalent genotypes (HPV-6/11/16/18/31/33/45/52/58) | 67 | 36.4 | 29.0 - 44.4 |
| Other than nonavalent genotypes | 69 | 34.9 | 27.8 - 42.7 |

Unadjusted PR of anal HPV (6/11/16/18 genotypes) comparing unvaccinated and vaccinated GBM was 1.4 (95%CI: 0.8-2.4). After adjustment for potential confounders and stratification by lifetime self-reported STI diagnosis (Appendix D), PR were 4.3 (95%CI: 1.2-13.8) and 0.6 (95%CI: 0.2-1.4) among GBM with and without a self-reported history of STIs, respectively. In the model, the interaction term between HPV vaccination and lifetime self-reported STI diagnosis was 7.5 (95%CI: 1.8-31.0).

### 3.7. Vaccination intention among unvaccinated GBM aged ≤26

Of 139 unvaccinated participants aged ≤26, 91 (65.4%) had heard of HPV vaccination. The RDS-adjusted proportion was 76.0% (95%CI: 66.6-83.4). Vaccination intention (within the next year) among unvaccinated GBM aged ≤26 aware of HPV vaccination was 60.0% (95%CI: 47.3-71.6). Table 4 presents the largest differences between GBM who intended to get vaccinated and those who did not or were undecided.

**Table 4.**
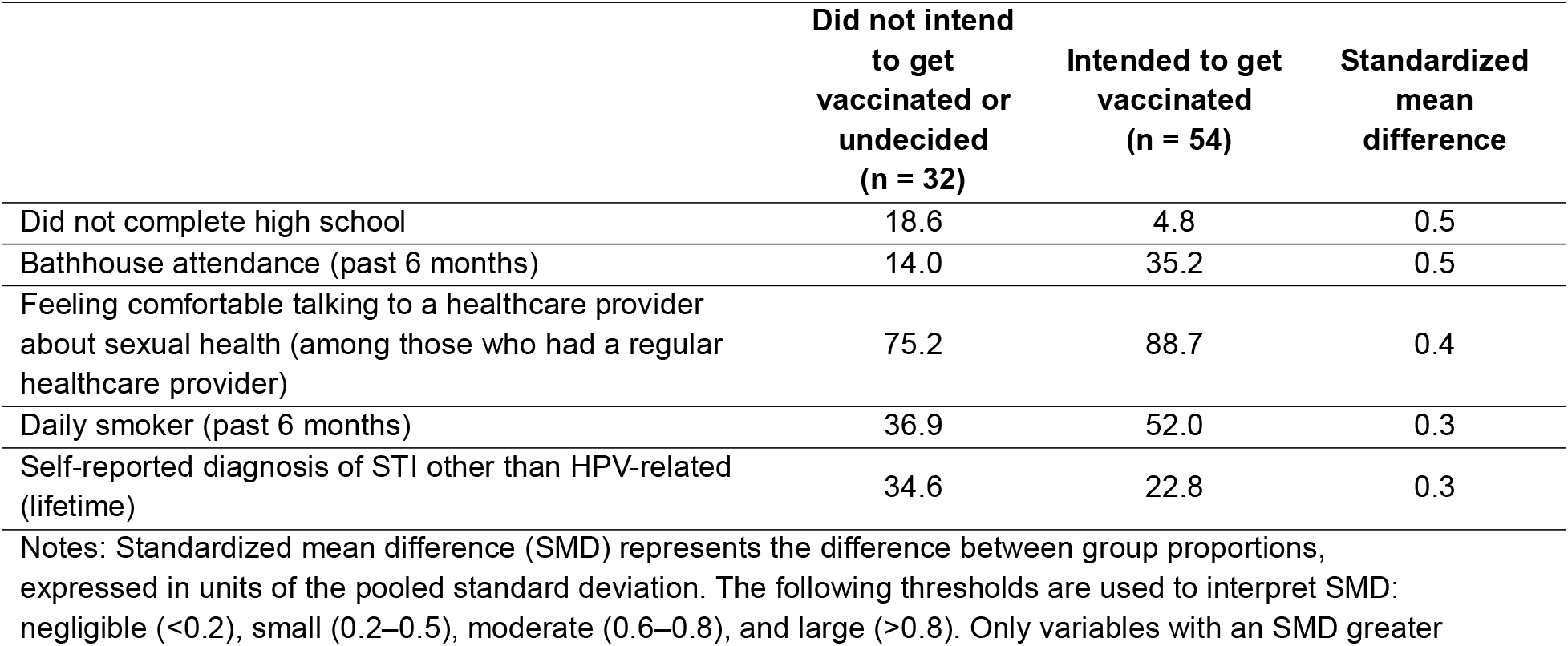
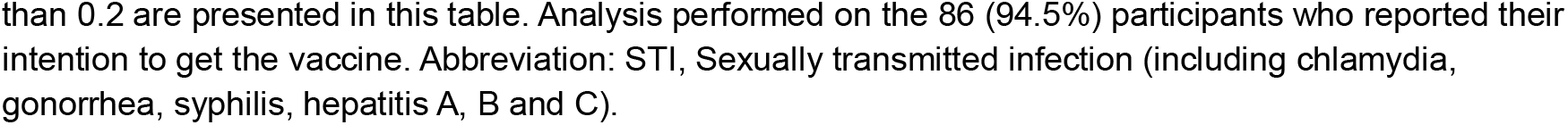
Largest differences between GBM who intended to get vaccinated and those who did not intend or were undecided, among unvaccinated GBM aged ≤ 26 years who had heard of HPV vaccination (n=86)

|  | Did not intend to get vaccinated or undecided (n = 32) | Intended to get vaccinated (n = 54) | Standardized mean difference |
| --- | --- | --- | --- |
| Did not complete high school | 18.6 | 4.8 | 0.5 |
| Bathhouse attendance (past 6 months) | 14.0 | 35.2 | 0.5 |
| Feeling comfortable talking to a healthcare provider about sexual health (among those who had a regular healthcare provider) | 75.2 | 88.7 | 0.4 |
| Daily smoker (past 6 months) | 36.9 | 52.0 | 0.3 |
| Self-reported diagnosis of STI other than HPV-related (lifetime) | 34.6 | 22.8 | 0.3 |

Notes: Standardized mean difference (SMD) represents the difference between group proportions, expressed in units of the pooled standard deviation. The following thresholds are used to interpret SMD: negligible ( $<0.2$ ), small (0.2–0.5), moderate (0.6–0.8), and large ( $>0.8$ ). Only variables with an SMD greater
than 0.2 are presented in this table. Analysis performed on the 86 (94.5%) participants who reported their intention to get the vaccine. Abbreviation: STI, Sexually transmitted infection (including chlamydia, gonorrhea, syphilis, hepatitis A, B and C).

## 4. DISCUSSION

### 4.1. Vaccination coverage

Over a year after Quebec extended its publicly-funded HPV vaccination to GBM aged ≤26, vaccination coverage (≥1 dose) reached 42% among eligible GBM in Greater Montreal, the province’s most populated region.

As reported in a previous Engage publication, vaccination coverage among GBM aged ≤26 (free vaccination age across Canada) was slightly higher in Montreal (35%) than in Toronto (33%) and Vancouver (26%)[13].

In our analysis, HPV vaccination was associated with previous hepatitis A or B vaccination, self-reported STI history, particularly genital warts, and in the past 6 months, HIV-Pre-Exposure Prophylaxis (PrEP) use and STI/HIV testing. Findings suggest that participants’ sexual activity possibly led to increased contact with the healthcare system, for example, through STI testing, during which participants could be offered HPV vaccination. This hypothesis aligns with vaccinated participants’ data showing 98% had already become sexually active before being vaccinated (Figure 1).

During Quebec’s 2023-2024 school year, 80% of boys aged 9-10 (grade 4) received ≥1 dose[26]. Engage study participants were ineligible for the school-based program extended to grade 4 boys (aged 9–10) in September 2016.

### 4.2. Disclosure of same-sex sexual activity to a healthcare professional

Only half of Montreal GBM eligible for free HPV vaccination had a regular healthcare provider. Therefore, relying solely on them to inform GBM about free HPV vaccination eligibility is insufficient. Relying on other healthcare providers (e.g., walk-in clinics, integrated STI prevention and screening services and community organizations) appears necessary to promote HPV vaccination among GBM.

Most participants who had a regular healthcare provider disclosed same-sex sexual activity to them before age 27 and 82% felt comfortable discussing sexual health with healthcare professionals, at an age young enough for HPV vaccination to be free and most effective.

Additionally, 82% of GBM eligible for free vaccination who both disclosed same-sex sexual activity and discussed HPV vaccination with their healthcare provider were vaccinated. Although unconfirmed, this suggests that many healthcare providers were aware of the publicly-funded HPV vaccination program for GBM.

### 4.3. Anal HPV Prevalence

Anal HPV prevalence among Montreal GBM eligible for free vaccination was 71% for ≥1 genotypes and 26% for ≥1 HPV-6/11/16/18 (quadrivalent-vaccine-genotypes). Anal HPV prevalence (6/11/16/18 genotypes) was not significantly lower among vaccinated GBM. Montreal Engage data and previous reports have shown that vaccinated GBM were more likely to engage in condomless receptive anal sex and to self-report a prior STI diagnosis than unvaccinated GBM, suggesting that targeted vaccination efforts are likely reaching those possibly previously HPV-exposed[13].

Clinical trials have shown 84% vaccine efficacy against incident HPV-6/11/16/18 genotypes among GBM who were HPV-naÏve, aged 16-26, had <5 lifetime sexual partners and no prior HPV or HIV history, consistent with expected vaccine efficacy among preadolescents[9].

Although emerging evidence suggests some benefit of catch-up vaccination, even with previous HPV exposure[11,27], vaccine effectiveness is expected to be lower in this real-world context. HPV vaccination is unapproved for therapeutic indications[28], and most men acquire ≥1 HPV infection within 2–4 years of sexual debut[29].

Furthermore, almost all vaccinated participants had sexual activity before getting vaccinated and half of them self-reported an STI history, therefore it is likely that several participants were infected with HPV-6/11/16/18 before vaccination. To maximize prophylactic benefits, school-based vaccination for preadolescents appears essential.

Anal HPV-6/11/16/18 prevalence among GBM aged ≤30 in Montreal (27%) approximated that in Toronto (29%) and Vancouver (23%)[14]. Likewise, genital HPV-6/11/16/18 prevalence among GBM aged 18-59 in the US (2013-2016) was 19%[30].

A previous Engage analysis which pooled Montreal, Toronto and Vancouver data showed an association between HPV vaccination (≥1 dose) and anal HPV-6/11/16/18 prevalence[14], while the analysis of Montreal data alone based on a smaller sample did not, which does not imply the absence of association in the target population. This variation between analyses may be explained by differences between cities, smaller overall statistical power (smaller sample size) and the focus of this analysis on eligible GBM for free vaccination. Additionally, self-reported vaccination status may lead to misclassification, potentially obscuring associations between vaccination and anal HPV prevalence[31] but not likely to meaningfully change the findings[32]. An Australian study showed that many young GBM misreported their HPV vaccination status by comparing participants’ self-reported vaccination status with medical records[33].

### 4.4. Vaccination intention among unvaccinated GBM aged ≤26

Among unvaccinated Montreal GBM aged ≤26, three-quarters had heard of HPV vaccination. Among these, 60% intended to get vaccinated within the next year, which is encouraging. Participants who intended to get vaccinated differed from undecided or unwilling participants by having higher risk sexual behaviours (more partners, more STIs, increased bathhouse attendance, etc.) and by greater vaccine benefits awareness.

### 4.5. Strengths and limitations

Study strengths include the use of RDS recruitment and the RDS weights to produce population-level inference. Additionally, the Engage study, as a well characterized cohort of GBM, provided a rich dataset to evaluate the HPV vaccination program for GBM.

Limitations include self-reported HPV vaccination status and the non-negligible number of non-received or invalid HPV specimens. However, the proportion of valid specimens resembled results from other studies among GBM using self-collection methods[34]. Self-reported HPV vaccination status has good sensitivity (92%) and moderate specificity (76%)[35] but may be associated with non-differential misclassification. Without knowing the temporality between HPV vaccination and infection acquisition further complicated detection of any association. Vaccination coverage and intentions may have changed since data collection (2017-2018). The COVID-19 pandemic and social distancing measures may also have impacted HPV vaccine coverage and prevalence among GBM. Finally, this analysis focused only on HPV prevalence and not on HPV persistence, the latter having more clinical relevance, particularly for HR-HPV.

## 5. CONCLUSION

Over a year after Quebec extended publicly-funded HPV vaccination to GBM aged ≤26, vaccination coverage among Montreal GBM eligible for free vaccination was 42%. It doubled (82%) among those who also disclosed same-sex sexual activity and discussed HPV vaccination with their healthcare provider. Although most GBM felt comfortable discussing their sexual practices with a healthcare professional before the age limit of 27, half of them did not have a regular healthcare provider. Therefore, it appears necessary to also rely on other resources to effectively promote HPV vaccination among GBM. Anal HPV prevalence, including vaccine-specific HPV-6/11/16/18 genotypes, was high among Montreal GBM eligible for free vaccination, highlighting the importance of continuing school-based vaccination efforts (before HPV exposure) to obtain maximum benefits. One quarter of unvaccinated GBM aged ≤26, had not heard of HPV vaccination, but among those who had, vaccination intention was high (60%), enhancing the importance of properly informing healthcare providers and GBM about the publicly-funded HPV vaccination program.

## Data Availability

Data will be made available upon reasonable request to the corresponding author.

## Acknowledgments

We would like to thank the Quebec Ministry of Health and Social Services for its financial support of this analysis. We also wish to thank the entire Engage-Montréal study team, the research staff, community engagement committee members, as well as their community partner agencies and the participants without whom the study and this analysis would not have been possible. We also thank the principal investigators of the Engage study and the Engage-HPV substudy (in alphabetical order): Ann Burchell, Joseph Cox, Alexandra de Pokomandy, Daniel Grace, Troy Grennan, Trevor A. Hart, Jody Jollimore, Nathan Lachowsky, Gilles Lambert, David Moore and Darrell HS Tan.

We also acknowledge the contribution of Julie Guenoun for conducting the HPV genotyping tests in Dr. François Coutlée’s laboratory at the CHUM. We thank Catharine Chambers and Ramandip Grewal for their assistance with data analysis. We are also grateful for Iulia Gabriela Ionescu’s help with manuscript revision.

## Data statement

Data will be made available upon reasonable request to the corresponding author.

