## Appendix A for "HPV prevalence, vaccination coverage and intention to get vaccinated among gay, bisexual, and other men who have sex with men: Evaluation of Quebec’s (Canada) HPV vaccination program"

**Appendix A** Key variables and corresponding Engage questionnaire items

| **Variables used in the analysis** | **Corresponding Engage questionnaire item** |
| --- | --- |
| HPV vaccination | |
| Vaccination status | Have you ever received one or more doses of the HPV vaccine?  *If no, the following question was skipped* |
| Age at vaccination (first dose) | How old were you when you received your first dose of the HPV vaccine? |
| Having heard of the HPV vaccine | Before today, had you ever heard of the HPV vaccine? |
| Having discussed the HPV vaccine | Before today, has any health professional ever discussed getting the HPV vaccine with you? |
| Intention to get vaccinated | The HPV vaccine is available free of charge for men (including trans men) who are 26 years of age or younger, who identify as gay, bisexual, or as a man who has sex with men, and who disclose this information to a healthcare professional.  Think about what you might do in the next year.  How likely or unlikely would you be to get the vaccine?  The possible answers were dichotomized as follows:   - Very unlikely, unlikely or undecided - Likely or very likely |
| Healthcare provider | |
| Having a regular healthcare provider | Do you currently have a regular primary health care provider, that is, someone you can go to for routine medical check-ups or for specific health concerns?  A regular primary health care provider can include, but is not limited to, a family doctor, a nurse practitioner, a walk-in clinic, or interdisciplinary health centre.  *If no, the following three questions were skipped* |
| Having disclosed same-sex sexual activity to a healthcare provider | Does your current regular primary healthcare provider know that you have sex with men?  *If no, the following two questions were skipped* |
| Age at which participants first disclosed same-sex sexual activity to a healthcare provider | How old were you the first time you told a primary healthcare provider that you have sex with men? |
| Feeling comfortable talking about sexual health issues with the healthcare provider | Do you feel comfortable talking to your current regular primary healthcare provider about health issues specific to being gay, bisexual, or a man who has sex with men? |
| Other key variables | |
| Self-reported diagnosis of sexually transmitted infection (lifetime) | Have you ever been told by a doctor or nurse that you had any of the following sexually transmitted infections? Chlamydia, Gonorrhea, Syphilis, Hepatitis A, Hepatitis B, Hepatitis C |
| Self-reported diagnosis of condylomas (lifetime) | Have you ever been told by a doctor or nurse that you had any of the following sexually transmitted infections? Genital or Anal Warts (Condylomas) |
