## Supplementary figures and images for "HPV prevalence, vaccination coverage and intention to get vaccinated among gay, bisexual, and other men who have sex with men: Evaluation of Quebec’s (Canada) HPV vaccination program"

### Appendix B

**Appendix B** Engage study participant flowchart

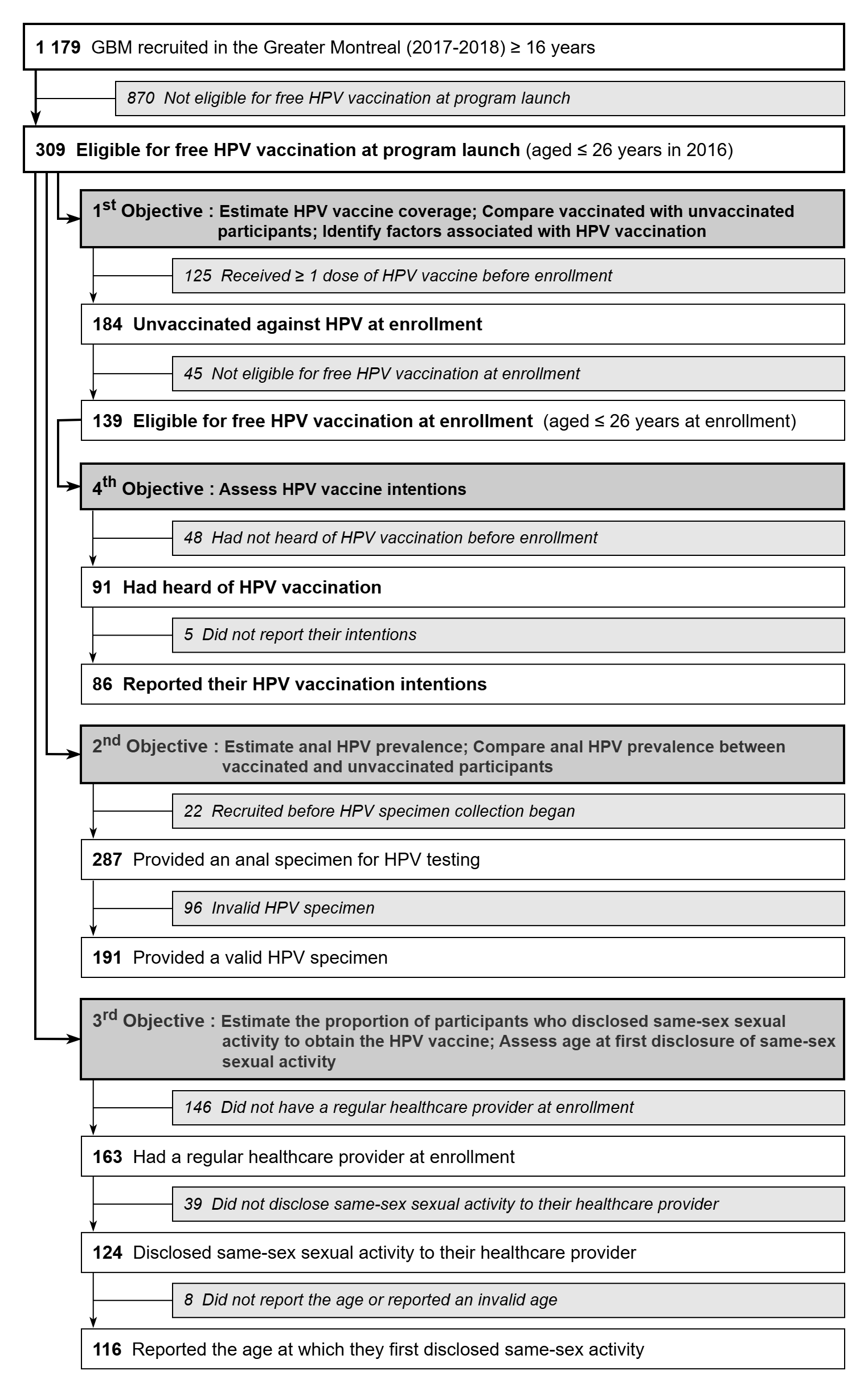
