## Appendix C for "HPV prevalence, vaccination coverage and intention to get vaccinated among gay, bisexual, and other men who have sex with men: Evaluation of Quebec’s (Canada) HPV vaccination program"

**Appendix C** Age of participants eligible for free HPV vaccination when they first disclosed same-sex sexual practices to their regular healthcare professional (n=124)

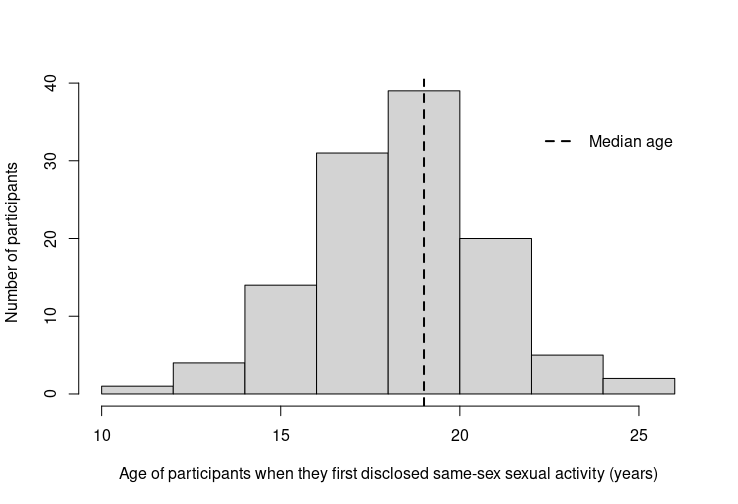
