## Appendix D for "HPV prevalence, vaccination coverage and intention to get vaccinated among gay, bisexual, and other men who have sex with men: Evaluation of Quebec’s (Canada) HPV vaccination program"

**Appendix D** Prevalence ratios (PR) of anal HPV infections (6/11/16/18 genotypes) comparing vaccinated (≥ 1 dose) with unvaccinated GBM eligible for free HPV vaccination (n=191)

|  | **Participants with anal HPV 6/11/16/18 genotypes** | | **Univariable regression** | | **Multivariable regression** | |
| --- | --- | --- | --- | --- | --- | --- |
|  | **Count** | **%** | **PR** | **95%CI** | **PR** | **95%CI** |
| Overall | | | | | | |
| Unvaccinated | 25 / 113 | 22.1 | Ref |  | Ref |  |
| ≥ 1 dose | 21 / 78 | 26.9 | 1.4 | 0.8 - 2.4 | 1.4 | 0.8 - 2.5 |
| Among participants who did not self-report diagnosis of STI (lifetime) | | | | | | |
| Unvaccinated | 22 / 81 | 27.2 | Ref |  | Ref |  |
| ≥ 1 dose | 5 / 25 | 20.0 | 0.6 | 0.2 - 1.7 | 0.6 | 0.2 - 1.4 |
| Among participants who self-reported diagnosis of STI (lifetime) | | | | | | |
| Unvaccinated | 3 / 32 | 9.4 | Ref |  | Ref |  |
| ≥ 1 dose | 16 / 53 | 30.2 | 4.7 | 1.4 - 16.2 | 4.3 | 1.3 - 13.8 |

STI: Sexually transmitted infection (including chlamydia, gonorrhea, syphilis, hepatitis A, B and C).
Notes: The multivariable regression models included the following potential confounders as covariate: age at enrollment ≤ 22 years, born in Canada, most comfortable speaking in French, education level, HIV status and HIV-PrEP use, daily smoking (past 6 months), ≥ 2 anal sex partners (past 6 months), and self-reported diagnosis of STI in the lifetime (first model only).
